# Declines in life expectancy following the COVID-19 pandemic in provinces of Spain

**DOI:** 10.1101/2021.04.15.21255545

**Authors:** Sergi Trias-Llimós, Amand Blanes, Manuel Franco, Usama Bilal, Tim Riffe

## Abstract

The COVID-19 pandemic has impacted population health on a global scale. Most of the studies on mortality impacts are at national level, while broad evidence exists on heterogeneous COVID-19 incidence across regions and within countries. Using Spanish data for 2020, we estimate life expectancy changes in 2020 compared with the 2017-19 period in 50 Spanish provinces. We visualize longer-term trends (1990-2020), and compare the robustness of our province-specific results with cumulative COVID-19 incidence using regional data from the Spanish ENECOVID seroprevalence study. In 2020 there was a 1.2 and 1.1 year drop in life expectancy for men and women in Spain, but this impact was heterogeneous across regions. For men these losses were highest in the province of Segovia (−3.5 years decline), while for women the highest drop was observed in Salamanca (−2.8 years decline). Life expectancy actually increased in Santa Cruz de Tenerife (+1.1 and +0.6 years for men and women, respectively). Declines in life expectancy in 2020 were also highly correlated with the cumulative seroprevalence through November 2020 (ρ=0.80 and 0.77 in men and women, respectively). Monitoring regional life expectancy dynamics provide valuable and granular information on the heterogeneous impacts of the pandemic on health at the population level. Similar exercises in other European countries may reveal insightful geographic patterns in mortality impacts in COVID-19 pandemic years.

## Introduction

The COVID-19 pandemic has impacted population health on a global scale (1,2). Spain has been one of the most severely affected countries within Europe, both in the first and in successive waves through 2020. Yet, the population-level impacts of COVID-19 in terms of mortality are challenging to assess due to differences in testing strategies and capacity, and differing definitions of infection across countries and over time, along with the concurrent direct and indirect effects of policies designed to mitigate the pandemic. Consequently excess mortality represents a more accurate measure of the population-level impact of COVID-19, as compared to laboratory confirmed COVID-19 mortality (3).

The most common and well-known mortality indicator used to capture mortality dynamics over time is life expectancy (see e.g. Luy et al., 2020; Trias-Llimós et al., 2020). Life expectancy represents the expected average lifespan of an individual if current mortality conditions were held fixed, a convenient and easily interpretable annualization of mortality rates. Recent estimates from Eurostat in April 2021 report that Spain showed the sharpest decline in life expectancy in the European Union between 2019 and 2020, with a loss of 1.6 years of life expectancy (6). Similar estimates for 29 countries worldwide report that Spain was one of the most affected countries in 2020, with 1.5 years of life expectancy drop between 2019 and 2020 (1). Such large declines had not been experienced in Spain since the Civil War, in the late 1930s (7).

These impacts have not been equally distributed across Spanish regions. In a previous study, we reported the heterogeneous impact of the first wave of the pandemic on life expectancy across Spanish NUTS-2 regions (*comunidades autonomas*), for example, life expectancy decreased around 2.1 (women) and 2.8 years (men) in Madrid, while very low impacts were observed in other regions including Canary islands, Murcia or Galicia (4).

In this study, we aimed to estimate life expectancy changes in 2020 compared with the 2017-19 period in 50 Spanish provinces with a complete calendar year of data for 2020 at a lower regional level, NUTS-3 level (*provincias*), accounting for all the waves contained in the first calendar year of the pandemic.

## Methods

We used mortality data from the “*Estimación del número de defunciones semanales (EDeS) durante el brote del covid-19”* from the Instituto Nacional de Estadística (INE), aggregated by 5-year age groups up to age 90+, gender, and province of occurrence (NUTS-3) for both 2019 and 2020. As the underlying electronic reporting system only captures 93% of deaths, the INE has corrected for this undercoverage using province-specific correction weights (8). We used populations as of July 1st, 2019 and 2020, as denominators to calculate mortality rates. To compare our results with longer term trends, we also obtained province-level life expectancy estimates for the 1990-2019 period.

We applied conventional life table techniques to estimate expectancy at birth by gender and province for both 2019 and 2020 (4). We compare our results with the estimates from INE for 2019, which confirmed that the use of provisional data corrected for coverage did not have a major impact on our life expectancy estimates. As life expectancy levels in small geographical areas can be affected by the randomness of the data in a given year we compared our 2020 estimates with the average for the period 2017-19 using INE estimates, and we show in the Supplementary Material (**Table S1**) the comparison between 2019 and 2020. We visualized the changes in life expectancy between 2020 and the average form 2017-19 for each province as well as historical trends (1990-2020).

Finally, we estimated the correlation between our estimated declines in life expectancy between 2017-19 and 2020 and the cumulative COVID-19 incidence across Spanish provinces by sex using cumulative seroprevalence data from the 4th wave of ENECOVID, a nationally representative seroprevalence survey (9,10). This indicator is a measure of the proportion of the population that tested positive for IgG SARS-CoV-2 antibodies in any of the four waves of the study.

Due to small population sizes, we excluded the Spanish autonomous cities of Ceuta and Melilla on mainland Africa from the analyses. Analyses were conducted in R version 4.0.2. Links to the original data sources can be found in Supplementary Material. Data and analysis code can be found at: https://osf.io/duqwx/

## Results

There was a 1.2 and 1.1 year drop in life expectancy for men and women in Spain in 2020 as compared to 2017-2019. Specifically, life expectancy in Spain fell from 80.6 to 79.4 years among men, and from 85.9 to 84.8 years among women (**Figure 1**). For men these losses were highest in the province of Segovia (−3.5 years), while for women the highest drop was observed in Salamanca (−2.8 years), both located in the Castille and Leon region. Life expectancy decreased the least, and actually increased, in Santa Cruz de Tenerife (+1.1 and +0.6 years for men and women, respectively). In four provinces, all belonging to the regions of Castilla-La Mancha and Castille and Leon in central Spain, the drops in life expectancy at birth were higher than 2.4 years in both men and women: Segovia, Salamanca, Ciudad Real, and Albacete. The two most populated provinces, Madrid and Barcelona, also experienced substantial declines in life expectancy: 2.8 and 1.9 among men and 2.2 and 2.0 among women for Madrid and Barcelona, respectively.

**Figure 1.**
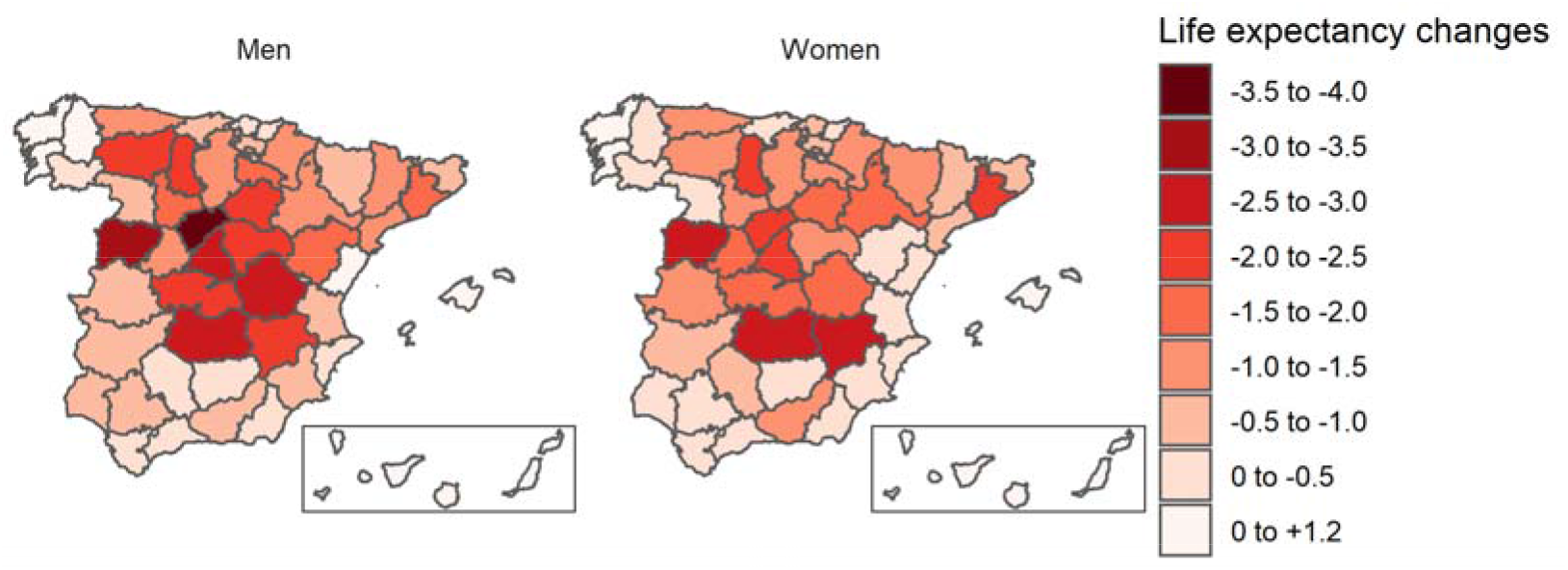
Life expectancy changes in Spanish provinces in 2020 as compared with 2017-19

The declines in the most affected provinces brought life expectancy back to levels seen 15-20 years ago (**Figure 2, Figure S1**). For example, among men and women, life expectancy in Segovia and Salamanca fell to the same levels as 2003/2004 and 1999/2000, respectively. For Madrid and Barcelona, the estimated life expectancy levels in 2020 resemble those in the years 2005-2008.

**Figure 2.**
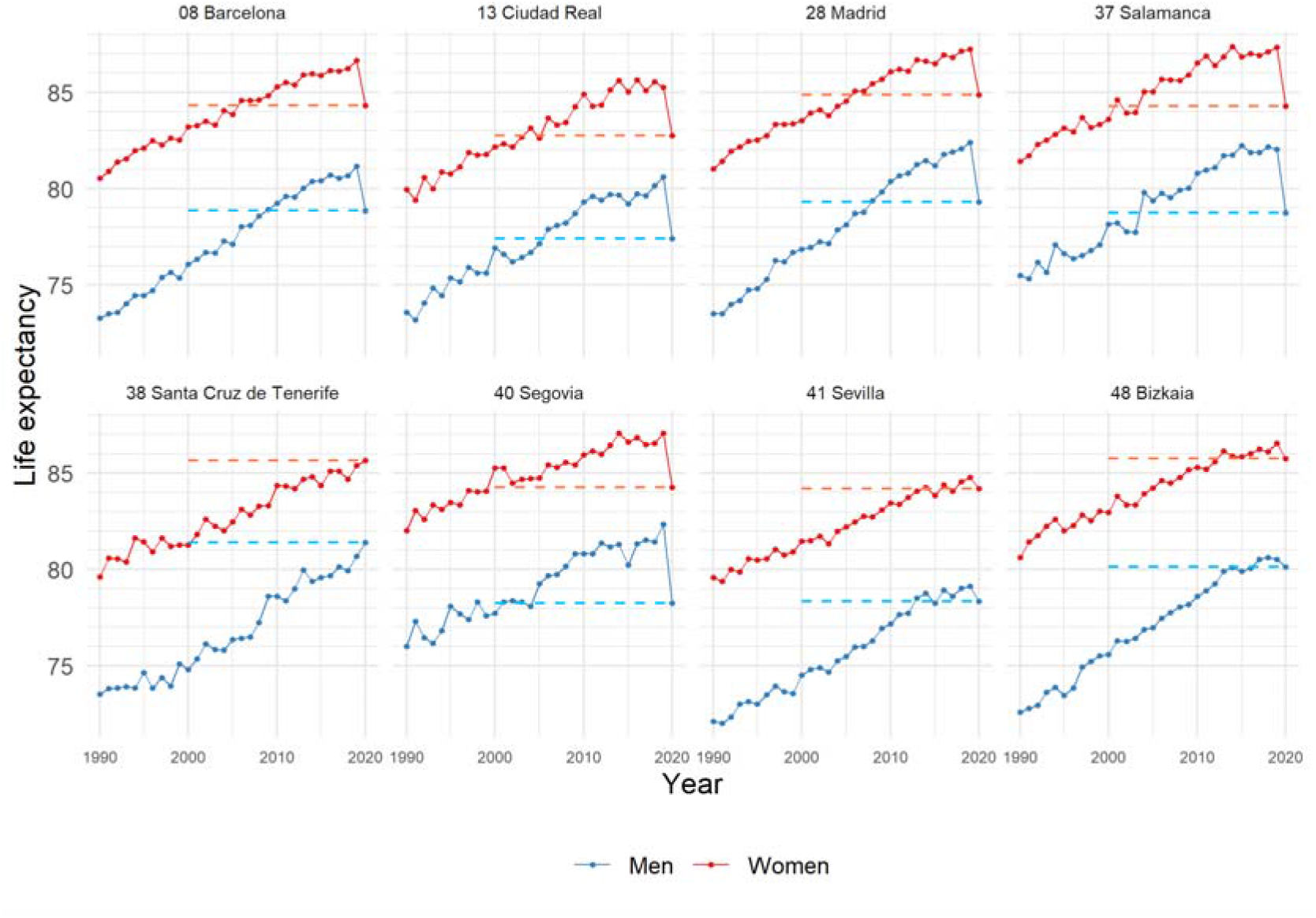
Life expectancy trends in selected Spanish provinces (1990-2020) *Results for the 50 Spanish provinces are presented in Figure S1.

Last, we found that changes in life expectancy were strongly correlated with SARS-CoV-2 seroprevalence through November 2020 (**Figure S2**, ρ=0.80 and 0.77 in men and women, respectively).

## Discussion

This study reported regional life expectancies for 2020 in Spain, the EU country that experienced the highest declines in life expectancy (6). Life expectancy changes between the period 2017-19 and 2020 were found to be heterogeneous across Spanish provinces with drops up to 3.5 years among men (Segovia) and 2.8 years among women (Salamanca).

This study used provisional data for 2020 on deaths reported by province of occurrence. The coverage of the original data was 93%, but the INE corrected for the province-specific undercoverage by applying weights. The data and the INE correction proves to be consistent and unbiased for 2019. That is, our life expectancy estimates using coverage-corrected estimates for 2019 are almost identical to those estimated by INE using final 2019 data (**Tables S1 and S2**). We should also acknowledge that, due to data limitations, the first age group used to estimate life tables was 0-4. In this case we assumed that the mean years of life lived by those dying in the interval (a0) was 0.5. This assumption implies a trivial effect on life expectancy estimates in low-mortality populations. Finally, we found a strong correlation with SARS-CoV-2 seroprevalence as of November 2020, lending validity to our findings.

This study reflects for the first time the extraordinary impacts that the COVID-19 has had on most of the Spanish provinces using conventional anual life expectancy estimates. The observed drops over 1 year of life expectancy between 2017-19 and 2020 in several regions as well as the national drops have not been experienced since the Civil War, in the late 1930s (7). In the first weeks of 2021, the third COVID-19 wave has also produced an observable excess mortality at the national level, but this impact differs from the one we reported here for 2020. For example, the provinces from the Valencian Community were highly affected, while its impact on 2020 was one of the smallest. This indicates that life expectancy for 2021 may not fully recover to the levels of previous years.

Monitoring regional life expectancy dynamics provide valuable and granular information on the heterogeneous impacts of the COVID-19 pandemic on health in different populations. Similar analyses in other countries may unreveal insightful geographical life expectancy dynamics.

## Data Availability

Links to the original data sources can be found in Supplementary Material. Data and analysis code can be found at: https://osf.io/duqwx/

## Declarations

## Availability of data and material

(data transparency): Links to original data can be found in Supplementary material

## Code availability

(software application or custom code): Data and analysis code can be found at: https://osf.io/duqwx/

## Authors’ contributions

(optional: please review the submission guidelines from the journal whether statements are mandatory): The study was conceived by STL and AB and designed by STL, AB, MF, UB and TR. STL analyzed the data with support from AB and input from UB and TR. STL drafted the main text with the support from UB, MF and TR. All authors approved the final manuscript and were responsible for the decision to submit for publication.

## Additional declarations for articles in life science journals that report the results of studies involving humans and/or animals

NA

## Ethics approval

(include appropriate approvals or waivers): NA

## Consent to participate

(include appropriate statements): NA

## Consent for publication

(include appropriate statements): The funders had no role in study design, data collection and analysis, decision to publish, or preparation of the manuscript.

