## Supplementary material for "Declines in life expectancy following the COVID-19 pandemic in provinces of Spain"

Links to the original data from Instituto Nacional de Estadística used in this study:

- Estimación del número de defunciones semanales (EDeS) durante el brote de covid-19: <https://www.ine.es/experimental/defunciones/experimental_defunciones.htm>
- Esperanza de Vida al Nacimiento por provincia, según sexo: <https://www.ine.es/jaxiT3/Tabla.htm?t=1485>
- Población residente por fecha, sexo y edad: https://www.ine.es/jaxiT3/Tabla.htm?t=9687&L=0

**Figure S1.** Life expectancy trends in Spanish provinces (1990-2020)

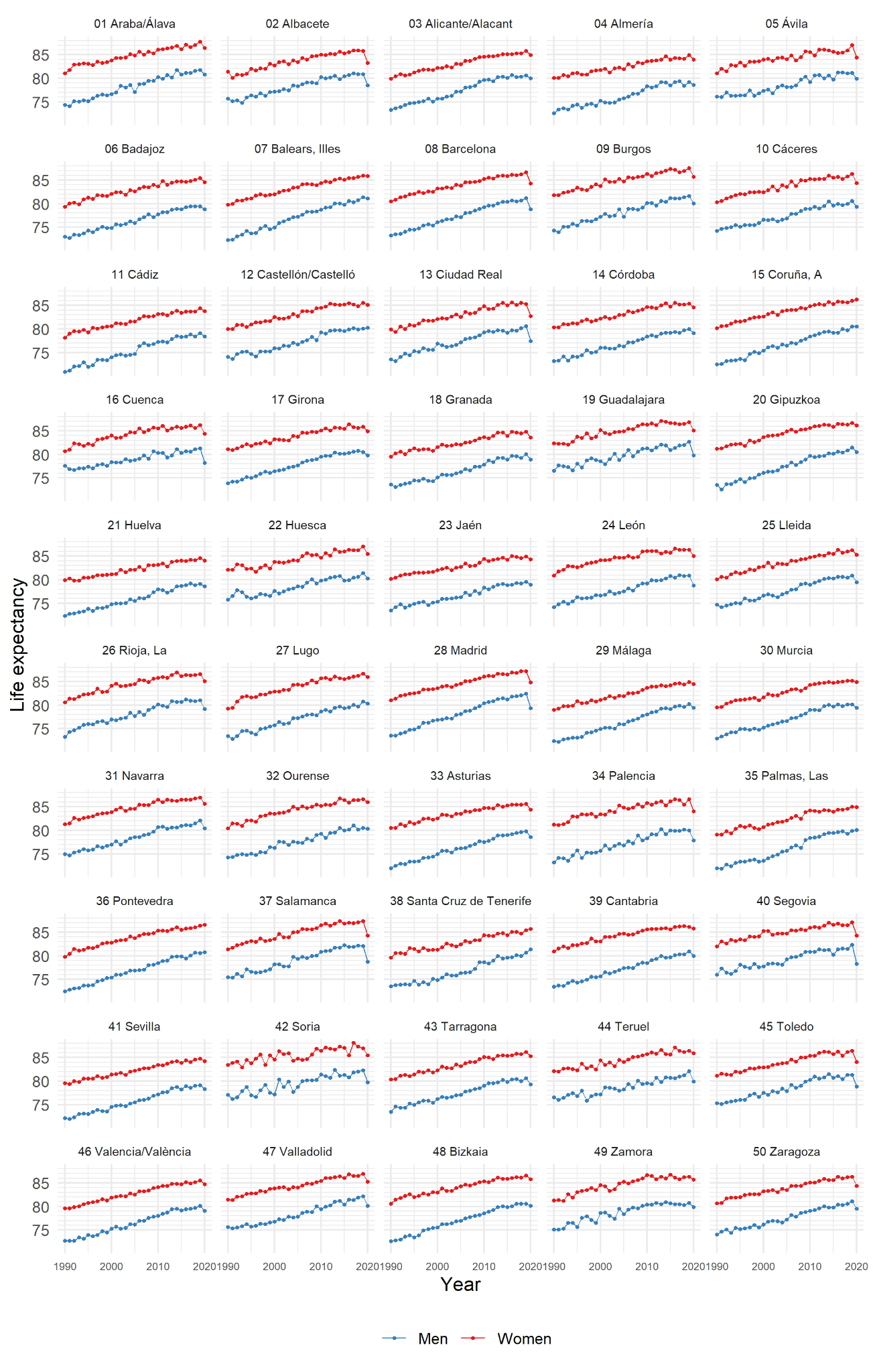

**Figure S2**. Associations between loss in annual life expectancy at birth between 2017-19 and 2020 and cumulative IgG anti SARS-Cov2 prevalence (4th round, December 2020) for Spainish provinces by sex

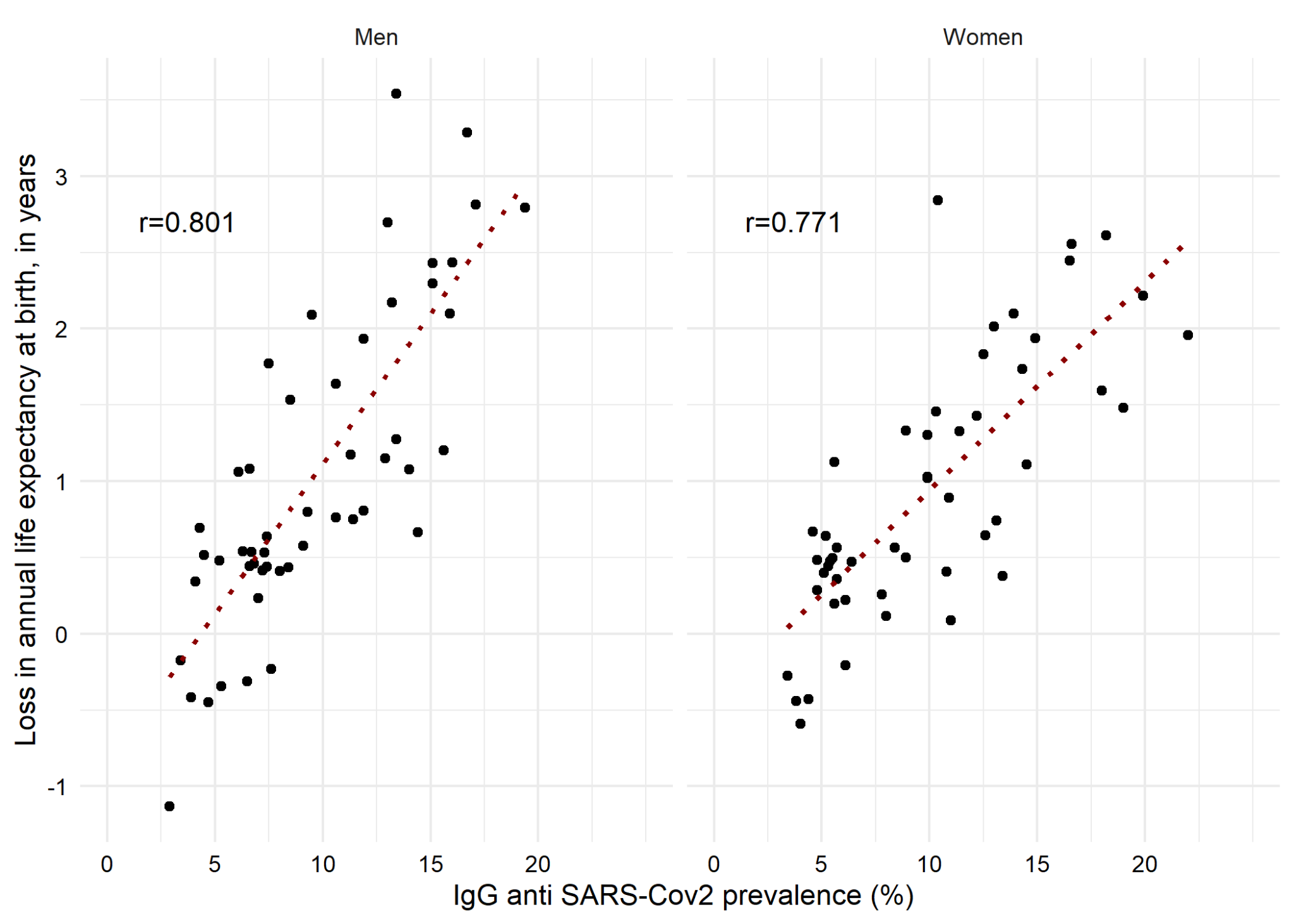

Note: Positive values in loss in life expectancy indicates a life expectancy drop, while negative values indicate a life expectancy gain.

**Table S1**. Life expectancy at birth estimates in 2020 and changes over time in Spanish provinces, men

|  | Own estimates | |  | INE estimates | | |  | Life expectancy changes | | |
| --- | --- | --- | --- | --- | --- | --- | --- | --- | --- | --- |
|  | 2019 | 2020 |  | 2019 | 2018 | 2017 |  | 2020-2019 | 2020-2019(INE) | 2020- (2017-19) |
| 01 Araba/Álava | 81.85 | 80.78 |  | 81.81 | 81.60 | 81.18 |  | -1.07 | -1.03 | -0.75 |
| 02 Albacete | 81.06 | 78.51 |  | 80.89 | 80.86 | 81.08 |  | -2.55 | -2.38 | -2.43 |
| 03 Alicante/Alacant | 80.94 | 79.99 |  | 80.66 | 80.35 | 80.29 |  | -0.95 | -0.67 | -0.44 |
| 04 Almería | 79.30 | 78.60 |  | 79.23 | 78.39 | 79.41 |  | -0.70 | -0.63 | -0.41 |
| 05 Ávila | 81.20 | 79.96 |  | 81.13 | 81.07 | 81.29 |  | -1.23 | -1.17 | -1.20 |
| 06 Badajoz | 79.58 | 78.81 |  | 79.48 | 79.46 | 79.41 |  | -0.76 | -0.67 | -0.64 |
| 07 Balears, Illes | 81.53 | 81.14 |  | 81.36 | 80.78 | 80.36 |  | -0.39 | -0.22 | 0.31 |
| 08 Barcelona | 81.27 | 78.86 |  | 81.16 | 80.68 | 80.53 |  | -2.42 | -2.30 | -1.93 |
| 09 Burgos | 81.84 | 80.09 |  | 81.63 | 81.36 | 81.09 |  | -1.75 | -1.54 | -1.27 |
| 10 Cáceres | 80.60 | 79.31 |  | 80.62 | 79.94 | 79.66 |  | -1.29 | -1.31 | -0.76 |
| 11 Cádiz | 79.20 | 78.43 |  | 79.10 | 78.40 | 78.82 |  | -0.77 | -0.67 | -0.34 |
| 12 Castellón/Castelló | 80.19 | 80.30 |  | 80.12 | 79.92 | 80.17 |  | 0.11 | 0.18 | 0.23 |
| 13 Ciudad Real | 80.69 | 77.43 |  | 80.60 | 80.15 | 79.63 |  | -3.26 | -3.17 | -2.70 |
| 14 Córdoba | 80.09 | 79.16 |  | 80.01 | 79.74 | 79.18 |  | -0.92 | -0.85 | -0.48 |
| 15 Coruña, A | 80.63 | 80.51 |  | 80.54 | 79.77 | 79.98 |  | -0.12 | -0.03 | 0.42 |
| 16 Cuenca | 81.25 | 78.17 |  | 81.25 | 81.07 | 80.56 |  | -3.08 | -3.08 | -2.79 |
| 17 Girona | 80.66 | 79.81 |  | 80.52 | 80.74 | 80.60 |  | -0.84 | -0.71 | -0.81 |
| 18 Granada | 80.24 | 78.86 |  | 80.05 | 79.30 | 79.64 |  | -1.38 | -1.19 | -0.80 |
| 19 Guadalajara | 82.92 | 79.76 |  | 82.70 | 82.00 | 81.88 |  | -3.15 | -2.94 | -2.43 |
| 20 Gipuzkoa | 81.54 | 80.48 |  | 81.45 | 80.83 | 80.40 |  | -1.06 | -0.97 | -0.41 |
| 21 Huelva | 79.19 | 78.57 |  | 79.12 | 78.88 | 79.25 |  | -0.63 | -0.55 | -0.52 |
| 22 Huesca | 81.48 | 80.28 |  | 81.42 | 80.61 | 80.41 |  | -1.20 | -1.14 | -0.53 |
| 23 Jaén | 79.63 | 78.93 |  | 79.56 | 79.22 | 79.32 |  | -0.70 | -0.63 | -0.44 |
| 24 León | 81.06 | 78.82 |  | 80.91 | 80.79 | 81.02 |  | -2.25 | -2.09 | -2.09 |
| 25 Lleida | 80.94 | 79.48 |  | 80.92 | 80.42 | 80.61 |  | -1.46 | -1.44 | -1.17 |
| 26 Rioja, La | 81.15 | 79.18 |  | 81.06 | 80.88 | 80.90 |  | -1.97 | -1.88 | -1.77 |
| 27 Lugo | 80.79 | 80.35 |  | 80.80 | 79.69 | 80.03 |  | -0.44 | -0.45 | 0.17 |
| 28 Madrid | 82.65 | 79.32 |  | 82.41 | 82.07 | 81.91 |  | -3.34 | -3.09 | -2.81 |
| 29 Málaga | 80.50 | 79.45 |  | 80.22 | 79.63 | 79.87 |  | -1.05 | -0.77 | -0.46 |
| 30 Murcia | 80.16 | 79.47 |  | 80.10 | 80.11 | 79.80 |  | -0.69 | -0.63 | -0.54 |
| 31 Navarra | 82.21 | 80.48 |  | 82.14 | 81.47 | 81.07 |  | -1.73 | -1.66 | -1.08 |
| 32 Ourense | 80.63 | 80.39 |  | 80.56 | 80.20 | 81.10 |  | -0.25 | -0.17 | -0.23 |
| 33 Asturias | 79.89 | 78.58 |  | 79.87 | 79.63 | 79.50 |  | -1.31 | -1.29 | -1.08 |
| 34 Palencia | 80.09 | 77.89 |  | 80.02 | 80.22 | 79.93 |  | -2.20 | -2.13 | -2.17 |
| 35 Palmas, Las | 80.15 | 80.14 |  | 79.94 | 79.26 | 79.86 |  | -0.02 | 0.20 | 0.45 |
| 36 Pontevedra | 80.78 | 80.78 |  | 80.62 | 80.64 | 80.04 |  | 0.00 | 0.16 | 0.34 |
| 37 Salamanca | 82.08 | 78.75 |  | 82.05 | 82.18 | 81.86 |  | -3.33 | -3.30 | -3.28 |
| 38 Santa Cruz de Tenerife | 81.23 | 81.39 |  | 80.70 | 79.95 | 80.14 |  | 0.17 | 0.69 | 1.13 |
| 39 Cantabria | 80.94 | 80.00 |  | 80.93 | 80.35 | 80.34 |  | -0.94 | -0.93 | -0.54 |
| 40 Segovia | 82.25 | 78.24 |  | 82.36 | 81.45 | 81.54 |  | -4.01 | -4.12 | -3.54 |
| 41 Sevilla | 79.26 | 78.35 |  | 79.13 | 79.03 | 78.62 |  | -0.90 | -0.78 | -0.57 |
| 42 Soria | 82.32 | 79.77 |  | 82.32 | 82.06 | 81.82 |  | -2.55 | -2.55 | -2.29 |
| 43 Tarragona | 80.75 | 79.31 |  | 80.64 | 80.01 | 80.46 |  | -1.44 | -1.33 | -1.06 |
| 44 Teruel | 82.07 | 79.90 |  | 82.08 | 81.25 | 80.96 |  | -2.18 | -2.18 | -1.53 |
| 45 Toledo | 81.40 | 78.90 |  | 81.29 | 81.29 | 80.41 |  | -2.51 | -2.39 | -2.10 |
| 46 Valencia/València | 80.22 | 79.09 |  | 80.12 | 79.69 | 79.54 |  | -1.13 | -1.03 | -0.69 |
| 47 Valladolid | 82.22 | 80.18 |  | 82.18 | 81.94 | 81.33 |  | -2.04 | -2.00 | -1.64 |
| 48 Bizkaia | 80.66 | 80.13 |  | 80.54 | 80.62 | 80.54 |  | -0.53 | -0.41 | -0.43 |
| 49 Zamora | 80.86 | 79.85 |  | 80.75 | 80.30 | 80.49 |  | -1.01 | -0.90 | -0.66 |
| 50 Zaragoza | 81.22 | 79.54 |  | 81.13 | 80.62 | 80.33 |  | -1.67 | -1.59 | -1.15 |
| 51 Ceuta | 78.82 | 78.17 |  | 78.54 | 78.63 | 76.65 |  | -0.65 | -0.37 | 0.23 |
| 52 Melilla | 78.90 | 78.18 |  | 78.24 | 78.23 | 77.82 |  | -0.72 | -0.06 | 0.09 |
| **Spain** | **80.91** | **79.37** |  | **80.86** | **80.46** | **80.37** |  | **-1.54** | **-1.49** | **-1.19** |

**Table S2**. Life expectancy at birth estimates in 2020 and changes over time in Spanish provinces, women

|  | Own estimates | |  | INE estimates | | |  | Life expectancy changes | | |
| --- | --- | --- | --- | --- | --- | --- | --- | --- | --- | --- |
|  | 2019 | 2020 |  | 2019 | 2018 | 2017 |  | 2020-2019 | 2020-2019(INE) | 2020- (2017-19) |
| 01 Araba/Álava | 88.17 | 86.49 |  | 87.74 | 87.05 | 86.62 |  | -1.68 | -1.25 | -0.65 |
| 02 Albacete | 85.90 | 83.29 |  | 85.82 | 85.95 | 85.92 |  | -2.62 | -2.53 | -2.61 |
| 03 Alicante/Alacant | 86.08 | 84.96 |  | 85.80 | 85.28 | 85.28 |  | -1.12 | -0.84 | -0.49 |
| 04 Almería | 85.14 | 84.02 |  | 84.93 | 84.19 | 84.28 |  | -1.12 | -0.91 | -0.44 |
| 05 Ávila | 87.21 | 84.42 |  | 87.03 | 85.96 | 85.47 |  | -2.79 | -2.61 | -1.74 |
| 06 Badajoz | 85.37 | 84.57 |  | 85.47 | 85.13 | 84.82 |  | -0.80 | -0.90 | -0.57 |
| 07 Balears, Illes | 86.19 | 85.91 |  | 85.99 | 85.67 | 85.44 |  | -0.28 | -0.08 | 0.21 |
| 08 Barcelona | 86.88 | 84.32 |  | 86.66 | 86.23 | 86.10 |  | -2.56 | -2.34 | -2.01 |
| 09 Burgos | 87.81 | 85.73 |  | 87.59 | 86.90 | 86.69 |  | -2.07 | -1.86 | -1.33 |
| 10 Cáceres | 86.37 | 84.34 |  | 86.29 | 85.81 | 85.29 |  | -2.03 | -1.95 | -1.45 |
| 11 Cádiz | 84.57 | 83.84 |  | 84.41 | 83.70 | 83.74 |  | -0.73 | -0.57 | -0.11 |
| 12 Castellón/Castelló | 85.72 | 85.14 |  | 85.53 | 84.88 | 85.26 |  | -0.58 | -0.39 | -0.09 |
| 13 Ciudad Real | 85.34 | 82.75 |  | 85.27 | 85.54 | 85.10 |  | -2.59 | -2.52 | -2.55 |
| 14 Córdoba | 85.47 | 84.63 |  | 85.43 | 85.22 | 85.25 |  | -0.84 | -0.80 | -0.67 |
| 15 Coruña, A | 86.30 | 86.26 |  | 86.04 | 85.67 | 85.74 |  | -0.05 | 0.22 | 0.44 |
| 16 Cuenca | 86.29 | 84.42 |  | 86.23 | 85.64 | 86.17 |  | -1.87 | -1.81 | -1.59 |
| 17 Girona | 86.13 | 84.88 |  | 85.92 | 85.63 | 85.76 |  | -1.25 | -1.04 | -0.89 |
| 18 Granada | 85.09 | 83.62 |  | 84.85 | 84.43 | 84.65 |  | -1.46 | -1.23 | -1.02 |
| 19 Guadalajara | 86.97 | 85.11 |  | 86.82 | 86.54 | 86.40 |  | -1.86 | -1.71 | -1.48 |
| 20 Gipuzkoa | 86.86 | 86.11 |  | 86.70 | 86.30 | 86.41 |  | -0.75 | -0.59 | -0.36 |
| 21 Huelva | 84.85 | 84.06 |  | 84.63 | 84.19 | 84.22 |  | -0.79 | -0.57 | -0.29 |
| 22 Huesca | 87.59 | 85.50 |  | 87.03 | 86.31 | 86.24 |  | -2.10 | -1.53 | -1.03 |
| 23 Jaén | 84.99 | 84.29 |  | 84.93 | 84.61 | 84.84 |  | -0.70 | -0.64 | -0.50 |
| 24 León | 86.63 | 85.06 |  | 86.34 | 86.38 | 86.36 |  | -1.57 | -1.28 | -1.30 |
| 25 Lleida | 86.27 | 85.29 |  | 86.30 | 86.05 | 85.74 |  | -0.98 | -1.01 | -0.74 |
| 26 Rioja, La | 86.78 | 85.11 |  | 86.60 | 86.41 | 86.32 |  | -1.67 | -1.49 | -1.33 |
| 27 Lugo | 86.86 | 85.94 |  | 86.67 | 86.26 | 86.08 |  | -0.92 | -0.73 | -0.40 |
| 28 Madrid | 87.65 | 84.86 |  | 87.24 | 87.16 | 86.81 |  | -2.79 | -2.38 | -2.21 |
| 29 Málaga | 85.24 | 84.45 |  | 84.94 | 84.39 | 84.67 |  | -0.80 | -0.49 | -0.22 |
| 30 Murcia | 85.18 | 84.92 |  | 85.16 | 85.17 | 85.03 |  | -0.26 | -0.24 | -0.20 |
| 31 Navarra | 87.14 | 85.67 |  | 86.95 | 86.83 | 86.56 |  | -1.47 | -1.28 | -1.11 |
| 32 Ourense | 86.77 | 86.02 |  | 86.63 | 86.45 | 86.41 |  | -0.75 | -0.61 | -0.47 |
| 33 Asturias | 85.80 | 84.42 |  | 85.63 | 85.49 | 85.51 |  | -1.39 | -1.21 | -1.12 |
| 34 Palencia | 86.72 | 84.10 |  | 86.64 | 85.46 | 86.49 |  | -2.62 | -2.54 | -2.10 |
| 35 Palmas, Las | 85.32 | 84.97 |  | 85.07 | 84.56 | 84.45 |  | -0.35 | -0.10 | 0.28 |
| 36 Pontevedra | 86.68 | 86.56 |  | 86.42 | 86.09 | 85.88 |  | -0.12 | 0.14 | 0.43 |
| 37 Salamanca | 87.54 | 84.28 |  | 87.34 | 87.12 | 86.92 |  | -3.26 | -3.06 | -2.84 |
| 38 Santa Cruz de Tenerife | 85.81 | 85.67 |  | 85.42 | 84.70 | 85.11 |  | -0.15 | 0.25 | 0.59 |
| 39 Cantabria | 86.33 | 85.74 |  | 86.14 | 86.29 | 86.22 |  | -0.59 | -0.40 | -0.47 |
| 40 Segovia | 87.47 | 84.25 |  | 87.07 | 86.53 | 86.49 |  | -3.22 | -2.82 | -2.44 |
| 41 Sevilla | 84.88 | 84.21 |  | 84.78 | 84.55 | 84.08 |  | -0.66 | -0.57 | -0.26 |
| 42 Soria | 87.20 | 85.50 |  | 87.01 | 87.29 | 88.08 |  | -1.70 | -1.51 | -1.96 |
| 43 Tarragona | 86.36 | 85.27 |  | 86.19 | 85.75 | 85.80 |  | -1.09 | -0.92 | -0.64 |
| 44 Teruel | 86.72 | 85.95 |  | 86.49 | 86.14 | 86.42 |  | -0.78 | -0.54 | -0.40 |
| 45 Toledo | 86.68 | 84.05 |  | 86.43 | 86.17 | 85.36 |  | -2.63 | -2.38 | -1.94 |
| 46 Valencia/València | 85.82 | 84.71 |  | 85.55 | 85.14 | 84.91 |  | -1.10 | -0.84 | -0.49 |
| 47 Valladolid | 87.31 | 85.23 |  | 86.96 | 86.50 | 86.52 |  | -2.08 | -1.73 | -1.43 |
| 48 Bizkaia | 86.73 | 85.75 |  | 86.55 | 86.12 | 86.26 |  | -0.99 | -0.80 | -0.56 |
| 49 Zamora | 86.38 | 85.72 |  | 86.27 | 86.23 | 85.80 |  | -0.66 | -0.55 | -0.38 |
| 50 Zaragoza | 86.61 | 84.33 |  | 86.34 | 86.20 | 85.95 |  | -2.28 | -2.01 | -1.83 |
| 51 Ceuta | 83.54 | 81.70 |  | 82.66 | 82.34 | 82.11 |  | -1.84 | -0.96 | -0.67 |
| 52 Melilla | 83.83 | 80.83 |  | 83.18 | 82.13 | 82.78 |  | -3.00 | -2.35 | -1.87 |
| **Spain** | **86.32** | **84.80** |  | **86.22** | **85.85** | **85.73** |  | **-1.52** | **-1.42** | **-1.13** |
